# Conditions for a second wave of COVID-19 due to interactions between disease dynamics and social processes

**DOI:** 10.1101/2020.05.22.20110502

**Authors:** Sansao A. Pedro, Frank T. Ndjomatchoua, Peter Jentsch, Jean M. Tchuenche, Madhur Anand, Chris T. Bauch

## Abstract

In May 2020, many jurisdictions around the world began lifting physical distancing restrictions against the spread of severe acute respiratory syndrome coronavirus 2 (SARS-CoV-2), giving rise to concerns about a possible second wave of coronavirus disease 2019 (COVID-19). These restrictions were imposed as a collective population response to the presence of COVID-19 in communities. However, lifting restrictions is also a population response to their socio-economic impacts, and is expected to increase COVID-19 cases, in turn. This suggests that the COVID-19 pandemic exemplifies a coupled behaviour-disease system. Here we develop a minimal mathematical model of the interaction between social support for school and workplace closure and the transmission dynamics of SARS-CoV-2. We find that a second wave of COVID-19 occurs across a broad range of plausible model input parameters, on account of instabilities generated by behaviour-disease interactions. We conclude that second waves of COVID-19–should they materialize–can be interpreted as the outcomes of nonlinear interactions between disease dynamics and population behaviour.

## Introduction

The COVID-19 pandemic has given rise to an “epidemic of models” [48]. Diverse mathematical models of SARS-CoV-2 transmission have been instrumental in capturing infection dynamics and informing public health control efforts to mitigate the COVID-19 pandemic and reduce the mortality rate. The concept of “flattening the curve” comes from model outputs that show how reducing the transmission rate through efforts such as contact tracing and physical distancing can lower and delay the epidemic peak [44].

On account of limited options for pharmaceutical interventions such as vaccines, and inadequate testing capacity in many jurisdictions, the COVID-19 pandemic has also been characterized by large-scale physical distancing efforts–including school and workplace closure–being adopted by entire populations despite heavy economic costs. Mathematical models of COVID-19 transmission and control show that physical distancing can mitigate the pandemic [24, 37, 44] and this has subsequently been backed up by empirical analyses of case notification data. These analyses show how mitigation measures have reduced the effective reproduction number of SARS-CoV-2 below one, meaning that each infected case infects less than one person on average [3, 8, 32]. However, the population’s willingness to support school and workplace closures could wane over time, as the economic costs of closure accumulate [6]. This has given rise to the possibility of a second wave of COVID-19 in many populations.

The large role played by physical distancing during the COVID-19 pandemic exemplifies a coupled behaviour-disease system, in which human behaviour influences infectious disease transmission and vice *versa* [4, 5, 11, 12, 31, 36]. These systems are part of a broader class of pervasive systems in which human behaviour both influences, and responds to, the dynamics of our environment. Hence, one might better speak of a single, coupled human-environment system, instead of just human systems or environmental systems in isolation from one another [17, 43, 45].

The interactions between disease dynamics and behavioral dynamics in COVID-19 are emphasized by research showing that the perceived risk of COVID-19 infection is a predictor of adherence to physical distancing measures [35] and moreover that individuals respond to the presence of COVID-19 cases in their population by increasing their physical distancing efforts [49]. In turn, physical distancing has been shown to reduce the number of cases [8], completing the loop of coupled behaviour-disease dynamics. Some models have already begun exploring this interaction between disease dynamics and individual behaviour and/or public health policy decision-making for COVID-19 [1, 23, 38, 39, 41, 50].

In particular, the social aspects of behaviour-disease interactions seem to be relevant for COVID-19 decision-making. Individuals do not necessarily make the best possible (most rational) response to the presence of COVID-19 cases in their population. Instead, it has been found that political leaders can be influential in convincing individuals to change their physical distancing efforts [1]. Additionally, jurisdictions experiencing outbreaks that start relatively late appear to learn from the experiences of jurisdictions that were affected earlier [42]. Meanwhile, other research emphasizes a need for more work on the socio-economic aspects of the pandemic [21]. These findings suggest that imitation and social learning processes are important for understanding interactions between disease dynamics and decision-making for COVID-19, which ultimately determine the fate of epidemic curve.

Here we model the coupled behaviour-disease dynamics of COVID-19 transmission and population support for school and workplace closure, using a simplified theoretical model. We opted for a simple model that avoids heterogeneities because our objective is to gain insights into potential interactions between social and behavioural dynamics. Public opinion evolves according to social learning rules [4, 43], and public opinion in support of closure depends both on COVID-19 case incidence and accumulated socio-economic losses due to school and workplace closure. A central decision-maker chooses a time to initially close schools and workplaces when the outbreak begins, but may subsequently open and close them again depending on how public opinion ebbs and flows. Meanwhile, disease dynamics are described by a compartmental epidemic model [18]. The details of our mathematical model are described in the Methods section. We analyze the model to characterize the conditions that give rise to a second wave of COVID-19 in the population.

## Results

### Mechanisms causing a second wave

At our baseline parameter values, time series of infection prevalence *I*(*t*) and support for closure *x*(*t*) exhibit nontrivial time evolution, including a second wave of COVID-19 infections (Figure 1a, b). These results illustrate the basic mechanisms underlying the model dynamics. As infection prevalence grows, support for closure rises and eventually crosses the 50% threshold by *t* = 80 days. After this, infection prevalence peaks and begins to decline. Support remains at a high plateau for a period of two months, after which support for closure wanes, causing restrictions to be lifted by *t* = 160 days. Shortly thereafter, prevalence begins to rise again. Support for closure correspondingly rises again, but not quickly enough to prevent a second wave of COVID-19 with a peak at t = 240 days that is higher than the peak of the first wave.

**Figure 1:**
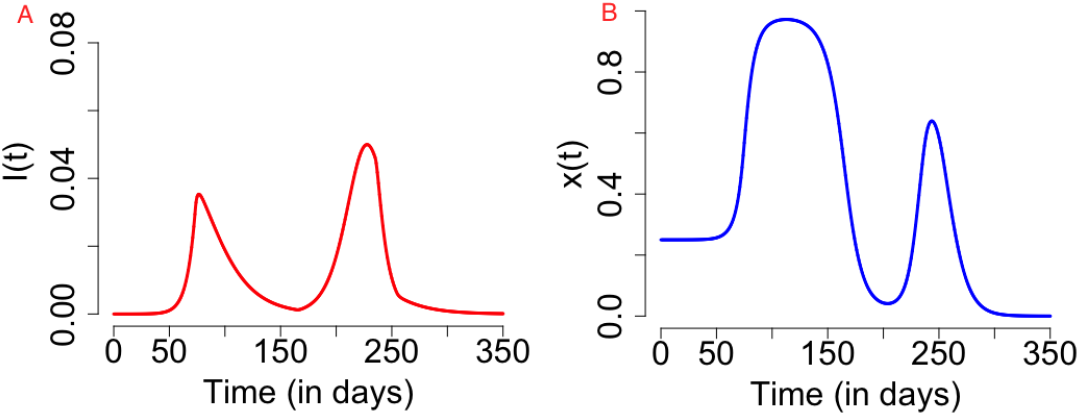
(left) Time series plot of the number of infected individuals in the face of social showing appearance of second wave (left) due to effects of social dynamics (right). The results are obtained for *C*_0_ = 0.6, *R*_0_ = 2.4,1/*σ* = 2.5, *γ* = 1/5, *κ* = 5, *ϵ* = 0.008, *α* = 1.0 * 24/365 and *δ* = 0.05 * 24.0/365 and initial conditions *S*(0) = (100000*α−*1)/1000000, *E*(0) = 0, *I*(0) = 1/1000000, *R*(0) = 0, *x*(0) = 0.25, *L*(0) = 0 and *H*(0) = 0.

### Epidemiological conditions for a second wave

Our goal was to gain insight into the conditions that generate a second wave of COVID-19, and to test the robustness of the predicted result at our baseline parameter values illustrate in Figure 1. Hence we explored the model dynamics in the neighbourhood of our baseline parameter values (Table 1) using parameter planes that show how the dynamical regimes of the model vary with changes in two model parameters (one on each axis) around the baseline values. We explored two dynamical outcomes: the number of waves in the course of the entire pandemic, and the ratio of the peak height of the second wave to the peak height of the first wave.

**Table 1:**
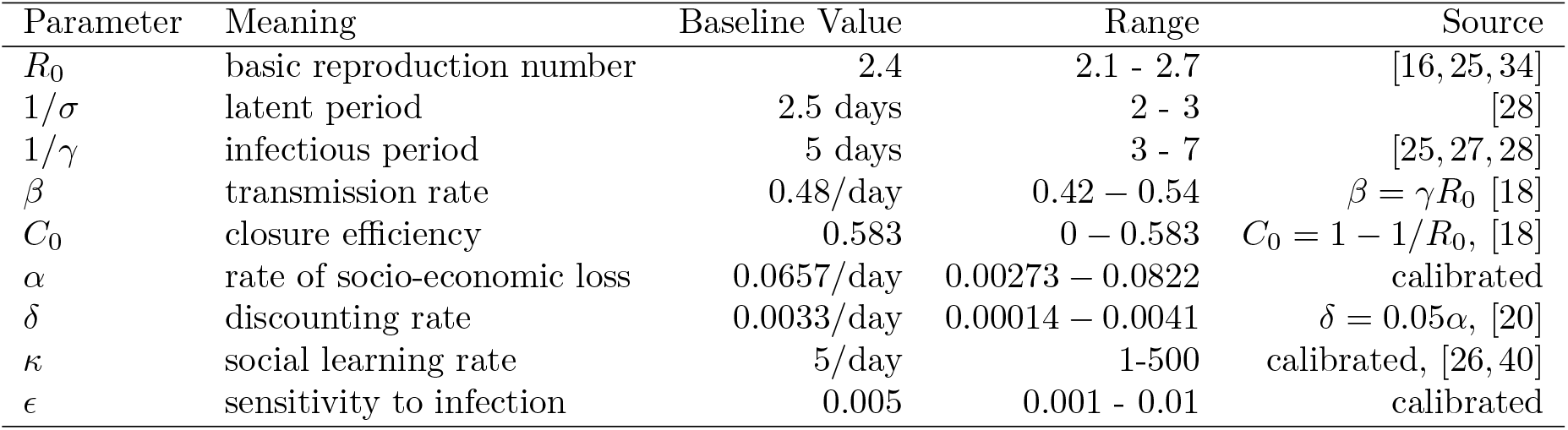
Parameter values, baseline values and literature sources

| Parameter | Meaning | Baseline Value | Range | Source |
| --- | --- | --- | --- | --- |
| $R_0$ | basic reproduction number | 2.4 | 2.1 - 2.7 | [16, 25, 34] |
| $1/\sigma$ | latent period | 2.5 days | 2 - 3 | [28] |
| $1/\gamma$ | infectious period | 5 days | 3 - 7 | [25, 27, 28] |
| $\beta$ | transmission rate | 0.48/day | 0.42 - 0.54 | $\beta = \gamma R_0$ [18] |
| $C_0$ | closure efficiency | 0.583 | 0 - 0.583 | $C_0 = 1 - 1/R_0$, [18] |
| $\alpha$ | rate of socio-economic loss | 0.0657/day | 0.00273 - 0.0822 | calibrated |
| $\delta$ | discounting rate | 0.0033/day | 0.00014 - 0.0041 | $\delta = 0.05\alpha$, [20] |
| $\kappa$ | social learning rate | 5/day | 1-500 | calibrated, [26, 40] |
| $\epsilon$ | sensitivity to infection | 0.005 | 0.001 - 0.01 | calibrated |

We start by exploring the effects of variation in the epidemiological parameters *β* (transmission rate) and *γ* (inverse of the average duration of infectiousness) in Figure 2, while keeping the rest of the parameters at baseline values. The results show that one, two, or three waves are possible under variation in these parameter values. A second wave characterizes most of the *β* − *γ* plane, however. A third wave appears when 1/*γ* is between 5 to 7 days and *β* is up to 0.4/day, corresponding to *R*_0_ *<* 2. (We note that most estimates place *R*_0_ *>* 2 for COVID-19 [19, 29].) The second peak may be higher or lower than the first peak, depending on the *β* and *γ* parameter combinations. For *R*_0_ *>* 2.4, the second peak tends to be lower than the first, while for *R*_0_ < 2.4 is is higher.

**Figure 2:**
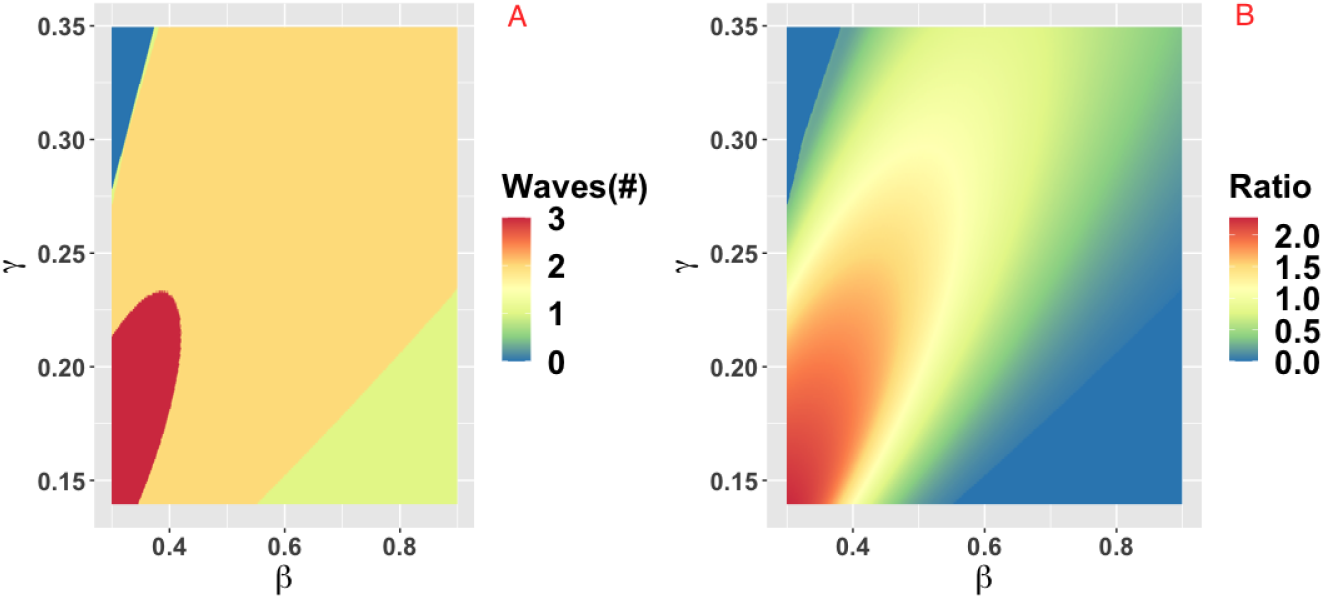
Parameter plane showing model dynamics as they vary with changes in the transmission rate *β* and the rate *γ* at which the infectious period ends, for the number of COVID-19 waves (a) and the ratio of peak height of the second wave to the first wave peak. Other parameters are at baseline values (Table 1).

Changes in the duration of infectiousness 1/*γ* and the duration of the latent stage 1/*σ* around baseline values do not change the number of peaks: a second wave is still observed across the range we explored (SI Appendix: Figure 2). However, the second peak is higher than the first when 1/γ is between 3 to 5 days, while out of this range the second peak is lower. The lack of dependence of dynamics on *σ* is expected. When 1/*γ* < 3 days, the second peak is less severe because *R*_0_ drops below levels that are feasible for continued transmission in the population. In contrast, when 1/*γ* > 5 days the second peak is less severe because a heightened *R*_0_ causes rapid build-up of herd immunity in the first wave of infection.

### Socio-economic and intervention conditions for a second wave

Next we explored the effect of variation in intervention, economic and social parameters. The parameter plane for *α*, the rate at which economic losses due to closure accumulate, and *δ*, the discounting rate for losses, shows little variation in these values across the ranges we explored (SI Appendix: Figure 3). Two waves are predicted and the peak of the second wave is higher than the first wave for almost all parameter combinations The only exception is that when *α* is very small, only a single wave occurs because the population is willing to tolerate economic losses indefinitely. As a result, *x* remains high over the entire time horizon of the simulation and COVID-19 is effectively controlled throughout this period.

The behavioural parameter *κ* is a measure of how quickly novel social behaviour spreads through a population as disease cases are reported. It has a large influence on the model dynamics, as represented in the *κ* − *α*, *κ* − *ϵ* and *κ* − *C*_0_ parameter planes (Figures 3-5). Higher values of *κ* indicate that individuals imitate more quickly. At our baseline value *κ* = 5/day, we observe a second wave. As the value of *κ* increases from this baseline value, the number of waves increases from two to six or seven in all three parameter planes, unless the effectiveness of closure (*C*_0_) is so low that the population experiences a single large epidemic that rapidly confers herd immunity to everyone (Figures 3-5). As *κ* is reduced sufficiently from its baseline value, the second wave is lost as expected, since we enter a parameter regime where the population responds with an unrealistic slowness to the presence of COVID-19, and it experiences a single, rapid pandemic wave that rapidly confers herd immunity. The second peak is higher than the first peak in all three parameter planes, except again when *C*_0_ is too low to effectively flatten the curve, and a single large outbreak results. Some examples of model outcomes for three or more waves are shown in SI Appendix: Figure 4. In these extreme scenarios, the second wave can either dominate the first and third waves, or it is also possible that the peaks of successive epidemic waves increase over time until it reaches a maximum peak in the fourth wave.

**Figure 3:**
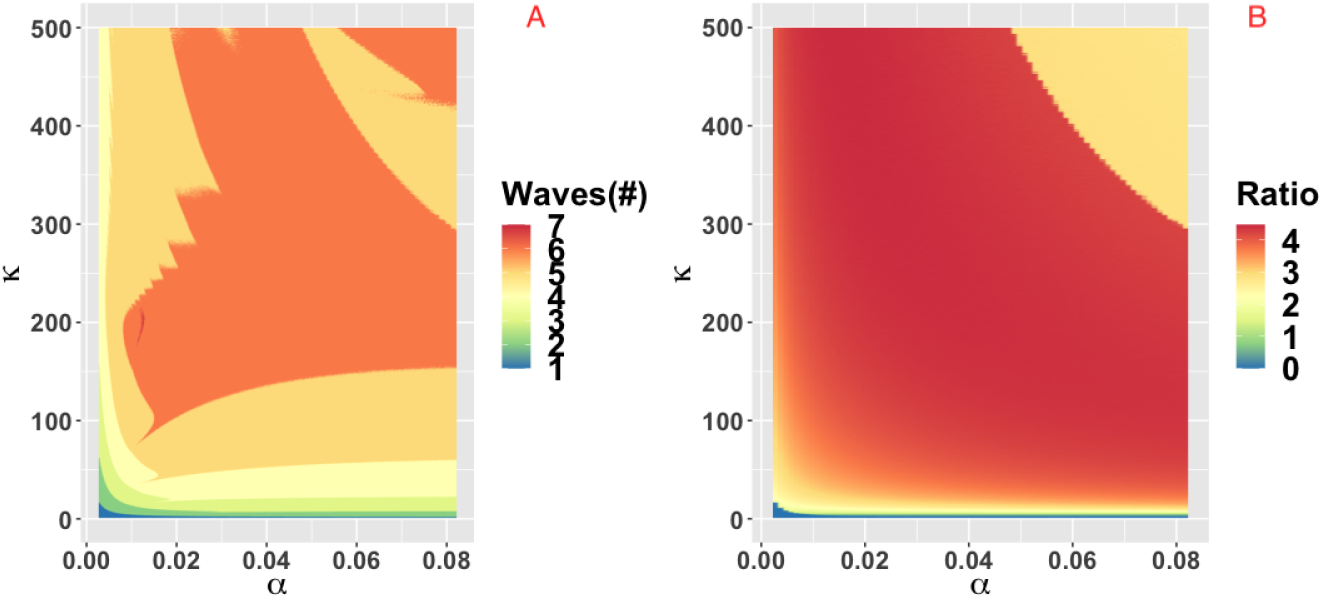
Parameter plane showing model dynamics as they vary with changes in the rate *α* at which socio-economic losses accrue and the rate *κ* that controls the social learning rate, for the number of COVID-19 waves (a) and the ratio of peak height of the second wave to the first wave peak. Other parameters are at baseline values (Table 1).

**Figure 4:**
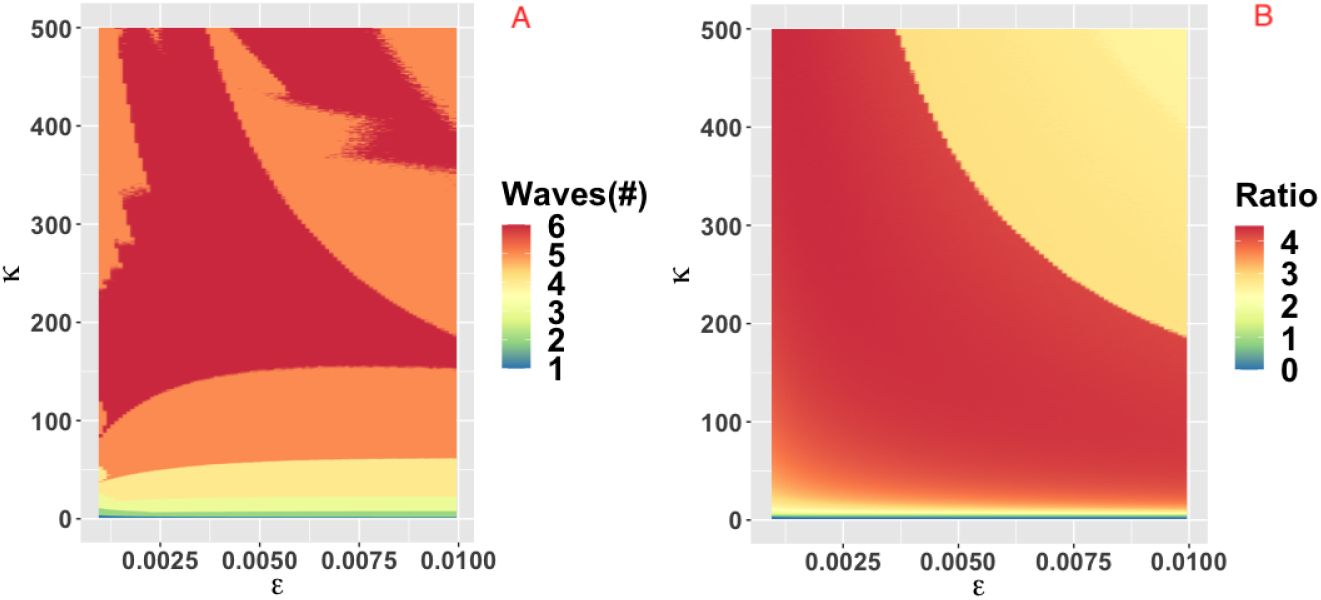
Parameter plane showing model dynamics as they vary with changes in the rate *κ* that controls the social learning rate and the parameter *ϵ* which controls how sensitive the population is to economic losses relative to infection prevalence, for the number of COVID-19 waves (a) and the ratio of peak height of the second wave to the first wave peak. Other parameters are at baseline values (Table 1).

**Figure 5:**
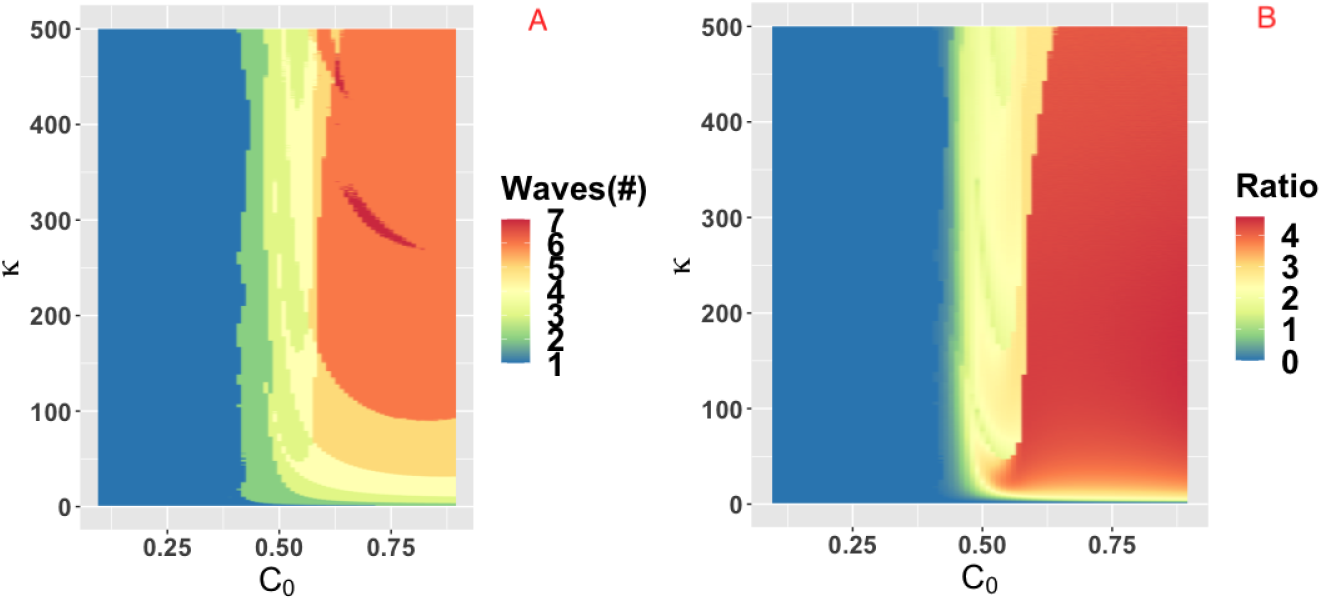
Parameter plane showing model dynamics as they vary with changes in the rate *κ* that controls the social learning rate and the parameter *C*_0_ which controls how effective closure is, for the number of COVID-19 waves (a) and the ratio of peak height of the second wave to the first wave peak. Other parameters are at baseline values (Table 1).

## Discussion

A second wave of COVID-19 is widely feared in May 2020 as many jurisdictions around the world begin lifting restrictions that have held viral transmission in check. To address this issue, we analyzed a simple theoretical model of the interplay between SARS-CoV-2 transmission dynamics and social dynamics concerning public support for physical distancing and school and workplace closure. We found that a second wave of COVID-19 (and sometimes also a third wave) was likely across a broad range of epidemiological and behavioural parameters. In some cases, the second peak was higher than the first peak, while for other parameter combinations it was lower.

Our prediction of a second wave driven by behaviour-disease interactions is plausible, given past and recent experience with novel emerging pathogens. One of the first affected countries in the COVID-19 pandemic–Iran–is now experiencing a large second wave on account of lifting restrictions in April 2020 [22]. During the 2003 SARS-CoV-1 epidemic in Toronto, premature relaxation of control measures resulted in a second wave of infections that was as large as the first wave [10]. Finally, behavioural responses to disease dynamics appear to have played a role in shaping the three waves that many populations experienced during the “Spanish flu” pandemic in 1918 [14].

Our model makes simplifying assumptions that could influence its projections. For instance, our model assumes that populations respond to infection prevalence *I*(*t*) but in fact, populations observe reported cases and deaths, both of which are delayed compared to time of actual infection. Time delays tend to destabilize dynamics in epidemic models [2] and hence we suspect that a model extension including a response to lagged outcomes like reported cases and deaths would exacerbate the severity of second waves in our model.

On the other hand, adding real-world spatial and demographic heterogeneities to our model could stabilize the dynamics and make the predicted oscillations less extreme, even if they do not remove them completely [9, 13, 30, 33, 46]. Similarly, on the behavioural modelling side, we suggest that the extreme oscillations observed in this model could also be stabilized if individuals use past and/or projected future states in their decision-making, instead of just the current prevalence, as we assumed [7, 47]. Alternatively, if individuals learn socially from other populations at differing stages of COVID-19 outbreaks [42], and not just their local population, this might also dampen the oscillations we observed in the model.

In summary, we speculate that incorporating social and spatial heterogeneities into the model would not completely remove the possibility of a second wave, although it could dampen the cycles [9, 13, 30, 33, 46] and give rise to epidemic curves more closely resembling that observed in the second wave in Iran [22]. Moreover, our prediction of a second wave was relatively robust across parameter space. Hence, we conclude that a second wave of COVID-19 on account of the coupled behaviour-diseases feedbacks we explore in this model will characterize many populations. We also conclude that that more effort in transmission modelling of COVID-19 should consider the effect of interactions between the dynamics of disease spread and social processes.

## Methods

### Model equations

Transmission dynamics are given by an SEIR model, modified to take physical distancing into account,

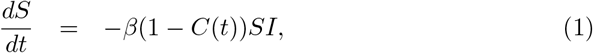

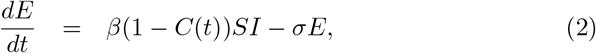

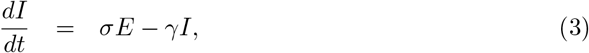

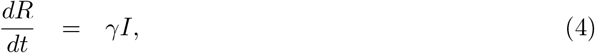

where *S* is the proportion of susceptible individuals (“susceptible”), *E* is the proportion of individuals who have been infected but are not yet infectious (“exposed”), *I* is the proportion of individuals who are both infected and infectious (“infectious”), and *R* is the proportion of individuals who are no longer infectious (“removed”). The time-varying parameter *C*(*t*) captures the impact of school and workplace closure on the transmission of COVID-19. *β* is the baseline transmission rate in the absence of school/workplace closure, *σ* is the time rate at which an exposed person becomes infections, and *γ* is the time rate at which an infectious person recovers.

The decision-maker decides to “turn on” closure at some time *t_close_*, and then decides to “turn off” closure when population support for closure, *x*(*t*) drops below 50%. Hence *C*(*t*) is given by:

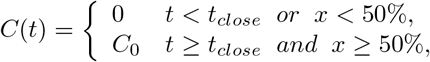

where *C*_0_ is a combined measure of how many workplaces are closed (the remainder being essential workplaces such as hospitals) as well as the effectiveness of physical distancing in those workplaces that remain open. The percentage of the population that supports school and workplace closure, *x*, evolves according to an imitation dynamic that represents social learning processes,

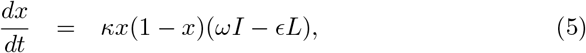

where *κ* is the social learning rate, *ω* is sensitivity to infection prevalence, and e is sensitivity to accumulated socio-economic losses *L*. In this equation, support for closure goes up when the prevalence of infection goes up, but it declines when the accumulated socio-economic losses, *L*, become too large. The quadratic term *x*(1 − *x*) represents a social learning dynamics where individuals sample others at some rate, and they change opinion based on the utility difference *ωI* − *ϵL*. (A full derivation of this type of differential equation appears in Ref. [43]). We can absorb *ω* into *κ*, yielding:

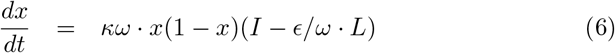

and then, setting *κ*′ = *κω* and *ϵ*′ = *ϵ*/*ω* and dropping the primes for simplicity we obtain

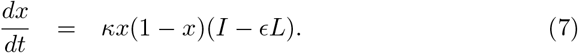

Finally, the variable *L* is a phenomenological representation of accumulated socio-economic losses obeying

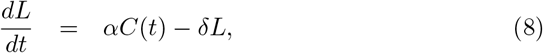

where *α* controls the rate at which school and workplace closure impacts socioeconomic health of the population, and *δ* is a decay rate that represents adjustment to baseline losses.

### Parameterization

A full list of parameter definitions, baseline values, and literature sources appears in Table 1. The basic reproduction number *R*_0_ and the transition rates *σ* and *γ*, were set based on COVID-19 epidemiological literature [16, 25, 27, 28, 34]. We note that the last compartment of the model, *R*, does not correspond to a stage of illness preceding recovery but rather a stage of infectiousness [2], which wanes quickly after the imposition of case isolation, in addition to the decline in viral shedding after the first 5 days [15]. Moreover, the infectious stages is preceded by a latent stage in which the virus is still replicating inside its new host until it can reach a level where the host can transmit the infection to others. These features of COVID-19 disease history guided our choice of *γ* and *σ*.

The social parameters *κ* and *ϵ* were calibrated. While *κ* dictates how quickly novel social behaviour spreads through a population, *ϵ* dictates how sensitive the population is to changes in case reports relative to socio-economic losses. *κ* can be estimated from behaviour early in the epidemic when socio-economic losses are small as *dx/dt* ≈ *κx*(1 − *x*)*I*. For *I*(*t*) we used reports of confirmed positive cases from the early stages of the United States epidemic, adjusted by a case under-ascertainment factor of 8.7 in the United States [26] in order to estimate the true prevalence of infections *I*(*t*) in the model. We used 21 January 2020 as the initial date of the epidemic, when the first case of COVID-19 was reported in the United States. Most populations rapidly adopted physical distancing measures against COVID-19. Gallup polls indicate that 59%/79%/92% of the United States public avoided going to events with large crowds, as of 13-15 March/16-18 March/20-22 March 2020 respectively [40]. Similarly, 30%/54%/72% avoided public places, and 23%/46%/68% avoided small gatherings [40]. Taking the average of these responses across the three question types, we obtain that *x*(52) = 0.373, *x*(59) = 0.597, *x*(66) = 0.773 where time is measured in days since January 21. Finally, we shifted these points forward 14 days as physical distancing at time *t* will not be reflected in infection data until *t* + 14 due to the delays in testing and symptom recognition. We assumed *x*(0) = 0.25 when fitting to these three data points using least-squares minimization for the *κ* calibration. Accordingly, we fit the model to the number of cases in the United States (SI Appendix: Figure 1b). The fitted infection trajectory is in good agreement with reported cases in the US during the initial phase of the epidemic leading up to 4 April 2020. For the special case where there is an absence of any control measures, the model predicts that about 80% of the population becomes infected by the end of the outbreak (SI Appendix: Figure 1a).

The remaining two parameters, *ϵ* and *α*, were calibrated to obtain the result that *x* remains high after the initial surge in support for closure, but begins to drop after two months. This period of time was based on the observation that two months that have elapsed since the declaration of the national emergency in the United States on 13 March 2020, and the process of re-opening state economies that has unfolded over the month of May 2020. These two parameters control the timescale of lifting school and workplace closure based on its socio-economic impacts. Finally, the parameter *δ* was set such that *δ* = 0.05*α* on the basis of commonly used discounting rates in economics and assuming that economic losses accumulated through the *αC*(*t*) term would be discounted at a rate of 5% per year [20].

In order to illustrate curve flattening and show that the model has the expected response to reduction in the transmission rate due to closure, we generated model timeseries of *I*(*t*) for the special case where closure is applied throughout the entire outbreak. The epidemic curve for different values of the closure efficacy *C*_0_ is shown, ranging from *C*_0_ = 0 (no intervention) to *C*_0_ = 0.6 (SI Appendix: Figure 1c). The timeseries show that the epidemic curve is flattened and delayed as closure becomes more efficacious, which reduces peak demand for intensive care beds and buys time for developing pharmaceutical interventions like vaccines and antiviral drugs, improving testing capacity, and establishing novel approaches to patient care. For the remainder of our analysis, to determine *C*_0_ it was assumed that *C*_0_ should be large enough to bring the effective reproduction number *R_eff_* below 1, reflecting the observed success in multiple jurisdictions where physical distancing and closure have maintained *R_eff_ <* 1 [3, 8, 32]. Hence we chose *C*_0_ = 1 − 1/*R*_0_ based on the elimination threshold for the SEIR model [18]. We also assumed *t_close_* = 20 days but in practice, our second requirement that *x* ≥ 50% was not reached until after 20 days in all of the model simulations.

## Data Availability

All data used to generate model projections are publicly available. The model simulation code is available upon request.

## Data availability

All data used in the model are publicly available.

### Computer code

The computer code used to generate our model projections is available upon request.

## Acknowledgements

This research was funded by NSERC Discovery Grants to CTB and MA.

